# *A* Progeroid Syndrome Caused *by RAF1* deficiency Underscores the importance of RTK signaling for Human Development

**DOI:** 10.1101/2022.02.20.22271260

**Authors:** Samantha Wong, Yu Xuan Tan, Kiat Yi Tan, Abigail Loh, Zainab Aziz, Engin Özkan, Hülya Kayserili, Nathalie Escande-Beillard, Bruno Reversade

## Abstract

Somatic and germline gain-of-function point mutations in RAF, the first oncogene to be discovered in humans, delineate a group of tumor-prone syndromes known as RASopathies. In this study, we document the first human phenotype resulting from the germline loss of function of the proto-oncogene *RAF1 (*a.k.a. *CRAF)*. In a consanguineous family, we uncovered a homozygous p.Thr543Met mutation segregating with a neonatal lethal progeroid syndrome with cutaneous, craniofacial, cardiac and limb anomalies. Structure-based prediction and functional tests using human knock-in cells showed that threonine 543 is essential to: 1) ensure RAF1’s stability and phosphorylation, 2) maintain its kinase activity towards substrates of the MAPK pathway and 3) protect from stress-induced apoptosis. When injected in *Xenopus* embryos mutant RAF1^T543M^ failed to phenocopy the effects of overactive FGF/MAPK signaling confirming its hypomorphic activity. Collectively, our data disclose the genetic and molecular etiology of a novel segmental progeroid syndrome which highlights the importance of RTK signaling for human development and homeostasis.

**Short summary:** A germline homozygous recessive loss-of-function mutation p.T453M in RAF1 causes a neonatal lethal progeroid syndrome. *In vitro* and *in vivo* tests demonstrate that Thr543 is necessary for RAF1’s protein stability, to transduce signaling to the MAPK pathway and to respond to stress-induced apoptosis.

## Introduction

Rapidly Accelerated Fibrosarcoma 1 (RAF1, MIM164760), is one of three mammalian RAF isozymes that delineate the conserved family of receptor tyrosine kinase (RTK) effectors in metazoans.^1^ This 70 kDA serine-threonine kinase is part of the Ras/Mitogen-activated kinase (MAPK) pathway. Binding of extracellular growth factors/mitogens to cell-surface RTKs (such as FGF/FGFR) activates RAS and subsequently RAF (ARAF (MIM311010), BRAF (MIM164757) and/or RAF1), triggering a phosphorylation cascade through MEK1/MEK2 and the ultimate effectors ERK1/ERK2.^2^ ERK1/ERK2 exert their function on numerous downstream nuclear and cytosolic targets culminating in increased cell proliferation. As such it is unsurprising that the earliest studies documented the somatic dysregulation of RAF1 as a primary driver of tumorigenesis.^1^ However, for the same reasons, more recent RAF research has revealed the regulatory role of RAF in physiological growth and most importantly in the context of this work, human embryogenesis.^2,3^ Many of these new insights have arisen from clinical observations of how germline mutations in regulators or components of the Ras/MAPK pathway exhibit a distinct but overlapping phenotype, owing to their common underlying mechanism.^2^

This class of congenital developmental disorders came to be referred to as “RASopathies”. It encompasses syndromes like Noonan syndrome (caused by mutations in *RAF1* (MIM611553), *PTPN11* (MIM151110), *SOS1* (MIM610733), *KRAS* (MIM609942) or *SHOC2* (MIM607721)), Costello Syndrome (*HRAS*, MIM218040), cardiofaciocutaneous syndrome (*BRAF, MAP2K1/2, KRAS*), neurofibromatosis type I (*NF1*, MIM162200), capillary malformation-arteriovenous malformation (*RASA1*, MIM139150), Noonan syndrome 13 (MAPK1, MIM176948) and Noonan syndrome 14 (SPRED2, MIM619745)^4^. Afflicted individuals often present with craniofacial dysmorphisms, ocular, musculoskeletal and cutaneous abnormalities, hypotonia, neurocognitive impairment, cardiac malformations and increased cancer risks.^2,5^ While these syndromes tend to be inherited in an autosomal dominant manner, many of them arise *de novo* in the parental germline or somatically in the patient and exhibit variable penetrance and severity.^6^

Of the approximately 22 genes implicated in RASopathies, those of the RAF family remain one of the best-studied owing to their cancer-associated roles. Genetic inactivations in mice have confirmed their non-redundant roles during development. *Araf*^-/-^pups die postnatally with distinct neurological and gastrointestinal abnormalities while *Braf*^-/-^ mice are embryonic lethal due to vascular defects.^7^ *Raf1*^-/-^ mice die by embryonic day e12.5, as a consequence of increased placental and liver apoptosis, while cardiac-specific *Raf1*^- /-^ mice exhibit extensive cardiac abnormalities.^8–10^ RAF1 is intricately controlled by numerous phosphorylation sites.^11^ In quiescent cells, phosphorylated Ser621 and Ser259 recruit 14-3-3, which enhances the binding of the inhibitory N-terminal CR1 domain of RAF1 to the catalytic C-terminal kinase domain. Upon mitogenic stimulation, activating residues such as Ser338, Ser289, Ser296, Ser301 and Ser642 are phosphorylated while inhibitory residues like Ser259 are dephosphorylated.^11,12^ In particular, RAF1 missense mutations have been associated with cardiac defects with p.S257L, p.P261T, p.L613V and p.T491I implicated in hypertrophic cardiomyopathy (HCM)^13^ and p.L603P, p.H626A, p.T641M and p.T310A in dilated cardiomyopathy (DCM).^14,15^ Consistent with their dominant mode of inheritance, the large majority of these detrimental RAF1 germline mutations exert gain-of-function activity vis-a-vis MAPK signaling. To date, no genetic data has been documented about the physiological requirement of wild type RAF1 during human development.

Here, we describe a previously unreported p.T543M variant in RAF1 segregating with a lethal progeroid phenotype in a consanguineous 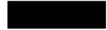 family. Afflicted neonates presented with split hand/foot malformations, craniofacial, cutaneous and cardiac abnormalities which are matching the commonly-affected organs seen in RASopathies. However, instead of the characteristic autosomal dominant inheritance, this phenotype was inherited in an autosomal recessive manner. Our collective *in vitro* and *in vivo* functional tests demonstrate that p.T543M behaves as a hypomorphic loss-of-function RAF1 mutant with impaired kinase activity, reduced protein half-life and increased cellular susceptibility to stress-induced apoptosis.

## Results

### Clinical phenotype and genetic analysis

Here, we report two affected siblings born to healthy, consanguineous first-cousin 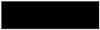 parents (Fig.1A). Both siblings, one male and one female, exhibited similar craniofacial, cardiac, and limb abnormalities suggestive for autosomal recessive inheritance. The first affected girl (III:1) had died on the seventh day of life due to necrotizing enterocolitis progressing to septicemia and no medical autopsy was available. The male proband was born via normal spontaneous delivery with a birth weight of 2670 g and length of 47.2 cm. He was hospitalized after birth for a week due to poor adaptation. He had craniofacial anomalies, complex cardiac anomalies, cleft hands and feet and was consulted at genetics outpatient clinics at 50 days of age. He had a weight of 1950 g, length of 51 cm and OFC of 33 cm. His general condition was very poor, hypotonic with no newborn reflexes. He had dry, lax, translucent skin with no subcutaneous fat. He had overriding cranial sutures with open frontal fontanelle of 2 × 2 cm, triangular face, prominent nasal root with hypoplastic alae nasi, narrow palpebral fissures, cleft palate, very small external ears with atresia of right external meatus, preauricular tag. He had split hands with missing 2^nd^ and 3^rd^ fingers, complete cutaneous syndactyly of 4-5^th^ fingers with fused nails on right and separate nail structures on left. The palmar lines were aberrant in both hands. There were 3 toes in each foot, 2^nd^ and 3^rd^ missing with cleft extending till distal metatarsal joints, total cutaneous syndactyly of 4-5^th^ toes and hypoplastic distal phalanx of hallux with rudimentary nail on one side and missing on the other. He had male genitalia and left cryptorchidism (Fig.1B). He had cardiac systolic murmur of 3/6 and cardiac ECHO revealed severe aortic stenosis, dysplastic aortic valve, muscular VSD, PDA;PFO and pulmonary hypertension and left ventricular dysfunction. Thin partial corpus callosum agenesis leading to wide occipital horns, colpocephaly, was present at cerebral USG. He had bilateral grade 3 hydroureteronephrosis and hypoplastic kidneys (R:31×17×19 and L:32×19×16 mm) with no corticomedullary differentiation. He died at 50 days of age due to cardiopulmonary arrest. No autopsy was performed. The aforementioned clinical features, albeit more severe, were reminiscent of Acro-Cardio-Facial Syndrome (ACFS, MIM600460), a congenital malformation syndrome with no identified genetic aetiology. Homozygosity mapping followed by exome sequencing revealed a unique germline homozygous missense mutation in *RAF1* (Chr:3 c.1628C>T, p.T543M) previously unrecorded in ExAC, UK10K, GnomAD or within our in-house exome databases (Fig. S1A). Sanger sequencing confirmed that this private *RAF1* mutation segregated with the disease in all available family members (Fig. S1B). Thr543 is highly conserved across vertebrate RAF1 orthologues (PhyloP=6.2, PhastCons=1) and RAF paralogues (Fig. S1A and Fig. 1D). It is situated in the kinase domain of RAF1 (Fig. 1C) and p.T543M is predicted deleterious in all databases (SIFT=0, PolyPhen=0.999, MCAP=0.818, PROVEAN=-4.12 and <u>MutationTaster</u>). Given RAF1’s role in transducing the growth-promoting activities of numerous RTKs during embryogenesis, we hypothesized that this homozygous germline RAF1^T543M^ allele might be the cause of this hitherto unknown progeroid syndrome.

**Figure 1.**
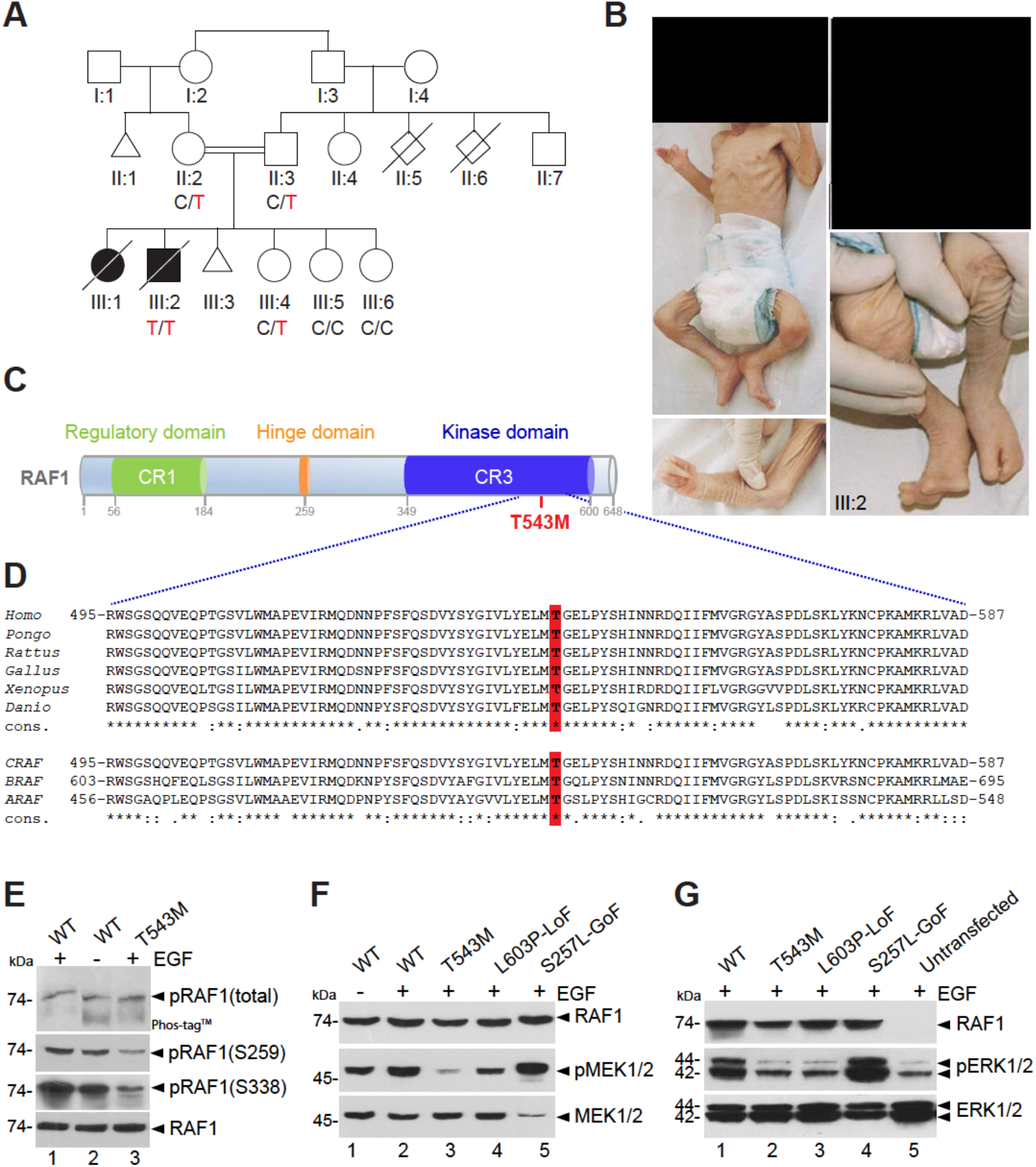
RAF1 germline homozygous missense mutation causes a neonatal lethal progeroid syndrome. **(A)** Pedigree of 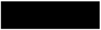 consanguineous family reporting the neonatal progeroid syndrome. Square, male; circle, female; black shading, affected individuals; small triangle, aborted; double lines, consanguineous marriages; diagonal line, deceased. **(B)** Pictures of affected proband (III:2) at 50 days showing general progeroid features.Note subcutaneous lipoatrophy, severe hypotonia, distinctive craniofacial features with absence of external ears (insets), cleft hands and feet with oligodactyly. **(C)** Schematic of the RAF1 protein structure showing the 3 conserved regions (CRs). p.T543M is located in the catalytic kinase domain (CR3). **(D)** Multiple sequence alignment showing conservation of T543 across species and among RAF family members. **(E)** RAF1^WT^ or RAF1^T543M^ were transiently overexpressed in HEK293T cells for 48 hours, followed by 16 hours of serum starvation and EGF stimulation for 15 min. Phos-tag™ gel showing global levels of RAF1 phosphorylation and corresponding western blot showing phosphorylation levels of Ser259 and Ser338 residues and total RAF1. **(F)** and **(G)** All the RAF1 constructs were transiently overexpressed in HEK293T cells as described in (E). Western blotting showing **(F)** phosphorylated MEK1/2, total MEK1/2 and total RAF1 and **(G)** phosphorylated ERK1/2, total ERK1/2 and total RAF1.

### The RAF1^T543M^ allele mimics a loss-of-function mutation in cultured cells

To investigate the pathogenicity of p.T543M, we assessed RAF1 and downstream MAPK/ERK activity in cultured cells (Fig. S1C). RAF1 activity is controlled by multiple phosphorylation sites.^11^ A Phos-tag® assay revealed that mutant RAF1^T543M^ remained globally hypophosphorylated (i.e. reduced electrophoretic mobility) compared to wild type RAF1 even if stimulated by exogenous EGF (Fig. 1E). Consistent with this, phospho-specific RAF1 antibodies against Ser259 and Ser338 showed a significantly lower signal for RAF1^T543M^ relative to RAF1^WT^ suggesting that it was refractory to EGF-triggered transduction (Fig. 1E). We next examined the MAPK/ERK pathway using three variants of RAF1 against which the RAF1^T543M^ can be evaluated: RAF1^WT^, a gain-of-function (GoF) allele RAF1^S257L^ (positive control), the most common activating mutation associated with HCM in Noonan syndrome and the heterozygous-acting loss-of-function (LoF) RAF1^L603P^ mutation associated with childhood-onset dilated cardiomyopathy (negative control).^14,15^ Western blot analyses showed that upon EGF stimulation, RAF1^T543M^ like the LoF construct RAF1^L603P^, failed to induce downstream MEK1/2 and ERK1/2 phosphorylation. Expectedly, RAF1^WT^, and even more so RAF1^S257L^, induced significantly more endogenous EGF-triggered MEK and ERK phosphorylation downstream of RAS/RAF1 (Fig. 1F and G, S1D). We conclude from these *in vitro* experiments that the p.T543M mutation brings about a loss-of-function in RAF1-mediated MEK/ERK signaling.

### The RAF1^T543M^ allele behaves as a loss-of-function mutation *in vivo*

We next assessed the signaling potential of these same RAF1 constructs in developing *Xenopus laevis* embryos, which provide a powerful *in vivo* test tube for FGF/MAPK signaling. Endogenous FGF/FGFR signaling is required for proper mesoderm and neural induction. Forced expression of dominant-negative or activating mutants of different components of this pathway yield tractable readouts.^16–19^ Upon overexpression in 2-to 4-cell stage embryos and western blots on extracts of stage 10 embryos showed that, like in HEK293T cells, the RAF1^T543M^ and the LoF RAF1^L603P^ causes significant reduced downstream ERK1/2 phosphorylation relative to the WT and GoF constructs (Fig. 2A and 2B). Phenotypically, increased RAF1^WT^ activity induced ectopic mesoderm differentiation. This effect was significantly augmented by GoF RAF1^S257L^ (Fig. 2C). This was readily quantified by increased *Xbra* expression in the circumferential layer of mesodermal cells at stage 10.5 and later by the development of supernumerary tails at stage 28 (Figure 2D and E). Like in HEK293T cells, RAF1^T543M^ induced minimal ectopic mesoderm which was comparable to that of LoF RAF1^L603P^ or uninjected controls (Fig. 2D and E). Similarly, N-tubulin staining of primary neuronal differentiation revealed that only overexpressed RAF1^wt^, but not RAF1^L603P^ or RAF1^T543M^ could induce ectopic neuronal differentiation, a hallmark of FGF-mediated posterization of the CNS (Fig. 2E). These results lend credence to the notion that the p.T543M allele is a hypomorphic mutation that cannot signal to its full context. Using gastrulating *Xenopus* embryos, we provide *in vivo* evidence that the missense p.T543M is a *bona fide* loss-of-function allele of RAF1 that cannot sustain FGF signaling.

**Figure 2.**
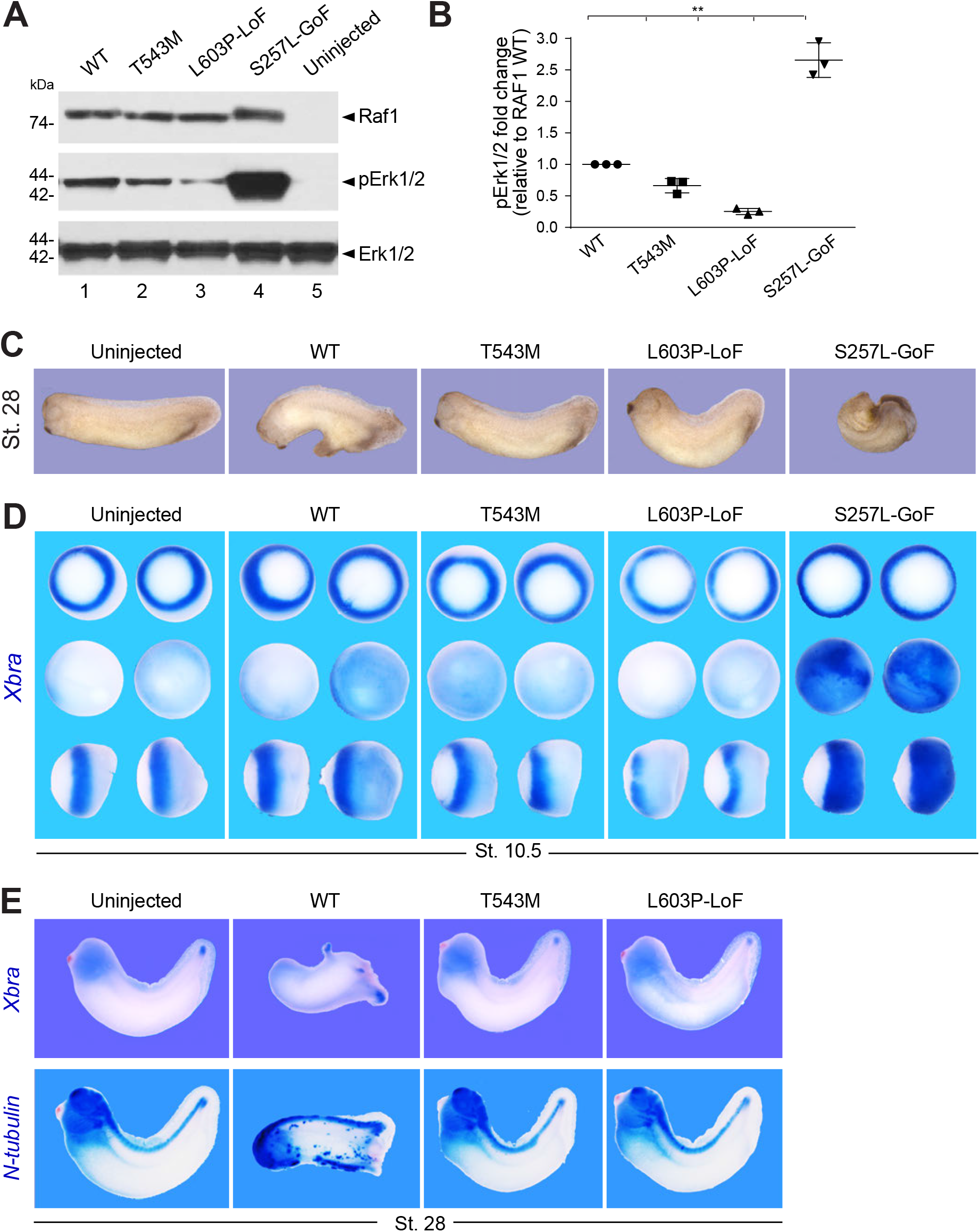
The RAF1^T543M^ allele behaves as a loss-of-function mutation *in vivo*. **(A)** Western blotting showing phosphorylated Erk1/2, total Erk1/2 and total Raf1 from *Xenopus laevis* embryos harvested at stage 28. **(B)** Quantification of phosphorylated Erk1/2, normalized to WT Raf1 in 3 independent western blots (Error bars indicate mean ± SEM. Two-tailed Student’s t test. *n*.*s*. not significant, **p<0.01). **(C)** Representative images illustrating the effect of WT or mutants *RAF1* mRNA injection into *Xenopus* embryos, at stage 28. **(D)** WISH performed on stage 10.5 *Xenopus* embryos injected with WT or mutants *RAF1* mRNA. *Xbra* (blue staining) marking mesoderm induction. Vegetal (top), animal (middle) and lateral (bottom) views. **(E)** WISH performed on stage 28 *Xenopus* embryos injected with WT or mutants *RAF1* mRNA. Blue staining reveals *Xbra* or *N-tubulin* markers depicting ectopic mesoderm induction, neural differentiation and brain posterization.

### Structure-based predictions and tests validate the pathogenicity of p.T543M

In silico structural analysis showed that Thr543 is located within the helical subdomain

∼22 Å away from the active site. Unlike Thr341 or Thr491, Thr543 is not known as a kinase-activating phosphorylation site (Fig. 3A). Instead, Thr543 is a polar residue mostly buried within the protein, with the beta-oxygen of its side-chain forming a hydrogen bond with main-chain Glu545 (Fig. 3B). A missense mutation to a methionine abolishes the hydrogen bond and causes steric clashes within the pocket (Fig. 3C). This can have knock-on effects towards either the active site or may deform a yet-unknown interaction site (Fig. 3C). Interestingly, the loss-of-function mutation p.L603P also affects a nearby helix located equidistant from the active site (Fig. S2B).^14^

**Figure 3.**
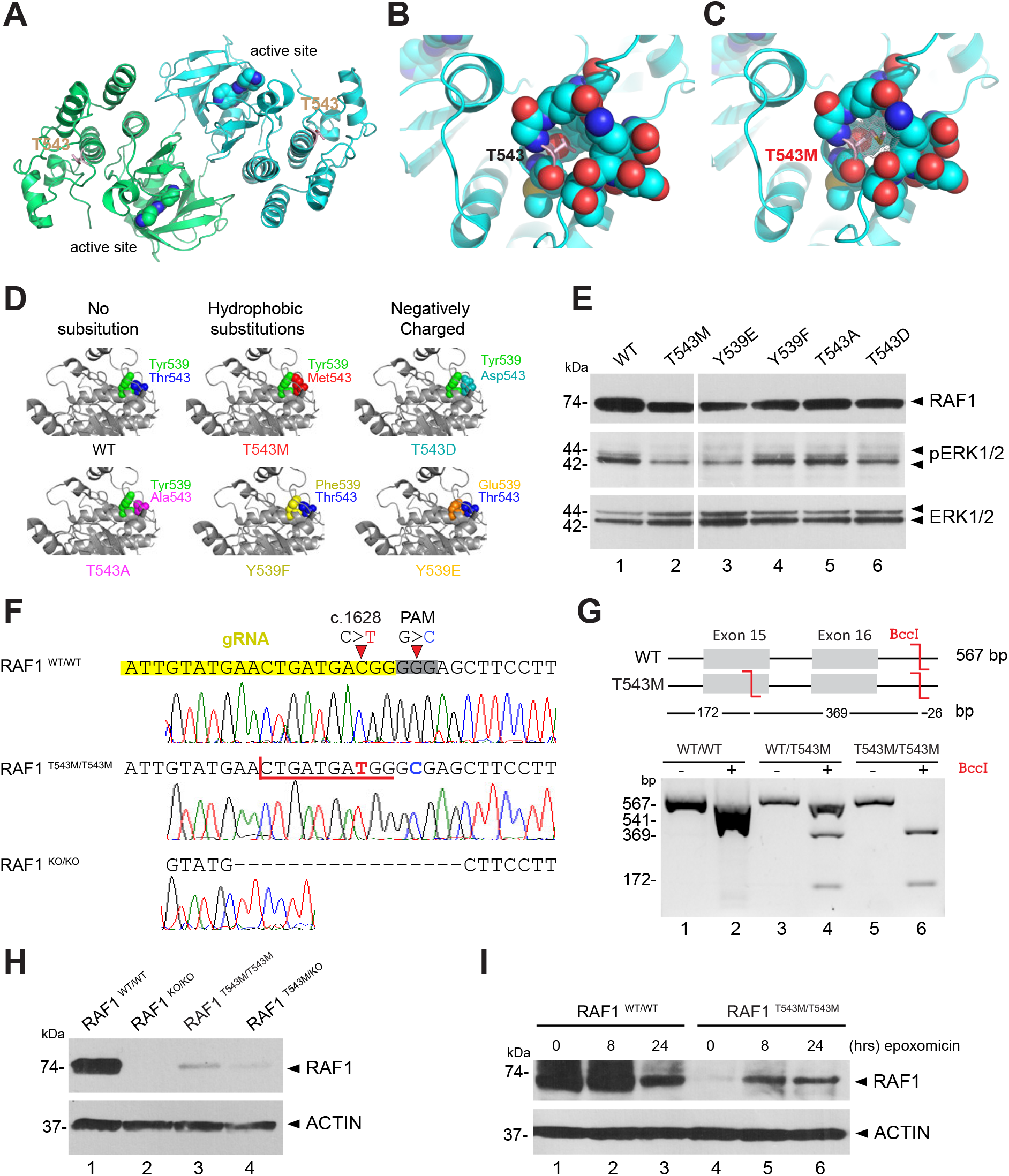
RAF1^T543M/T543M^ knock-in cells reveal RAF1’s compromised protein half-life. **(A)** RAF1 homodimer (PDB:3OMV) showing the location of T543 (drawn as ball-and-stick) on helical subdomain. Active site is occupied by an inhibitor drawn as spheres. **(B)** Positioning of Thr543 versus **(C)** Met543 in the pocket. The Met543 side chain clashes despite being modeled in the least sterically hindered rotamer. **(D)** Predicted side chain interaction of Thr543 and Tyr539 variants. **(E)** Western blot downstream signaling potential of structural mutants that induce steric (p.T543M) or electrostatic (p.Y539E or p.T543D) interference. **(F)** RAF1^T543M^ and RAF1^KO/KO^ HEK293T cells were generated by CRISPR-Cas9. Top - bottom: Sanger-sequencing of RAF1 in WT HEK293T cells; a RAF1^T543M^ clone showing the desired c.1628C>T (p.T543M) mutation and a synonymous PAM-disrupting c.1632G>C ; a RAF1^KO/KO^ clone with a homozygous 17bp deletion. **(G)** from gDNA of the genotypes shown with BccI. c.1628C>T introduces a BccI restriction site. The RAF1^WT/T543M^ heterozygote genotype was obtained from the patient’s father. **(H)** Western Blot for endogenous RAF1 in HEK293T cell lines of genotypes indicated, at steady state. **(I)** Western-blot showing that 8h and 24h of epoxomicin treatment is sufficient to partially rescue RAF1 in RAF1^T543M/T543M^ cells. ACTIN serves as loading control.

To investigate the predicted importance of Thr543 on local RAF1 structure, we substituted Thr543 and Tyr539, which closely packs onto Thr543 (Fig. 3D) (PDB: 3OMV ^20^), with a range of other amino acids *in silico*. Of these, p.T543A and p.Y539F (substitutions with shortened side chains) still fitted within the pocket and retained local structure and RAF1 activity (Fig. 3D). Conversely, we also modeled substitutions that likely disrupted RAF1 structure and kinase activity via steric (p.T543M), electrostatic means p.Y539E, or both p.T543D (Fig. 3D). These predictions were verified experimentally in HEK293T cells. Substitutions that shortened the side chains (p.Y539F and p.T543A) preserved RAF1 kinase activity (Fig. 3E), as measured by downstream phosphorylation of ERK1/2, whereas substitutions that likely cause steric (p.T543M) or electrostatic (p.Y539E and p.T543D) disruptions indeed led to a loss of kinase activity (Fig. 3E). These results suggest p.T543M abrogates RAF1 kinase activity potentially through disruption of a crucial local and possibly global structure within its kinase domain. As p.T543A and p.Y539F are incapable of phosphorylation, these results also argue against the involvement of a possible phosphorylation at these sites to control RAF1 function.

### Knock-in cells reveal a reduced half-life for endogenous RAF1^T543M^

While we demonstrated that overexpressed RAF1^T543M^ is kinase-deficient, several reports have found that the kinase activity is required for RAF1’s inherent stability by blocking its proteasomal degradation.^21,22^ To investigate this phenomenon, we generated a biallelic *RAF1* p.T543M knock-in HEK293T line using CRISPR/Cas9-mediated homology-directed repair (HDR) (Fig. 3F). Due to the incomplete nature of HDR, we also obtained complete RAF1^KO/KO^ carrying a homozygous 17-bp deletion (p.Tyr539AlafsTer26) and compound heterozygous RAF1^T543M/KO^ lines. Clonality of the homozygous RAF1^T543M/T543M^ line was confirmed at the genomic level through restriction digestion with endonuclease *BccI* (Fig. 3G). Compared to the WT parental line, western blotting for endogenous RAF1 showed drastically reduced protein levels that mirrored their respective genotypes. Homozygous RAF1^T543M/T543M^ cells displayed no more than 10-20% of wildtype RAF1 levels (Fig. 3H) while RAF1^T543M/KO^ cells had a further reduction consistent with having only one copy of mutant RAF1 being expressed. Endogenous RAF1^T543M^ protein half-life could be partially rescued following an 8-hour treatment with epoxomicin (Fig. 3I), arguing that RAF1^T543M^ is inherently unstable and targeted for proteasome-mediated degradation. This is in agreement with past reports showing that the kinase activity of RAF1 is essential for its proteostasis.^21^ Our results indicate that the p.T543M mutation not only diminishes RAF1’s kinase activity but also compromises its own protein half-life.

### RAF1^T543M/T543M^ knock-in cells are more sensitive to stress-induced apoptosis

RAF1 promotes cellular survival by antagonizing apoptosis through MEK/ERK-independent pathways.^23,24^ Hence, we next examined whether RAF1^T543M/T543M^ cells were more sensitive to apoptotic stimuli. Hydrogen peroxide (H2O2) was applied to mutant and isogenic control 293T cells and levels of apoptosis were measured using SYTOX™ green nucleic acid stain^25^. Fluorescent images showed that H2O2-treated RAF1^T543M/T543M^ cells displayed a greater number of SYTOX™-positive green nuclei compared to RAF1^WT/WT^ cells (Fig. 4A). We observed a dose-dependent and statistically significant difference in cell death (from 7% to 34%) in RAF1^T543M/T543M^ cells compared to parental wildtype cells (Fig. 4B). These findings suggest that beyond impaired MEK/ERK signaling which is expected to blunt cellular proliferation, the homozygous RAF1^T543M/T543M^ variant heightens cellular sensitivity to stress-induced apoptosis. It is noteworthy that ASK1 and MST2 are pro-apoptotic molecules that are directly antagonized by wild type RAF1 (Fig. 4C);^10,13,26,27^ hence we suspect that the lack of repression by RAF1^T543M/T543M^ may explain the observed increase in stress-induced apoptosis in knock-in 293T cells.

**Figure 4.**
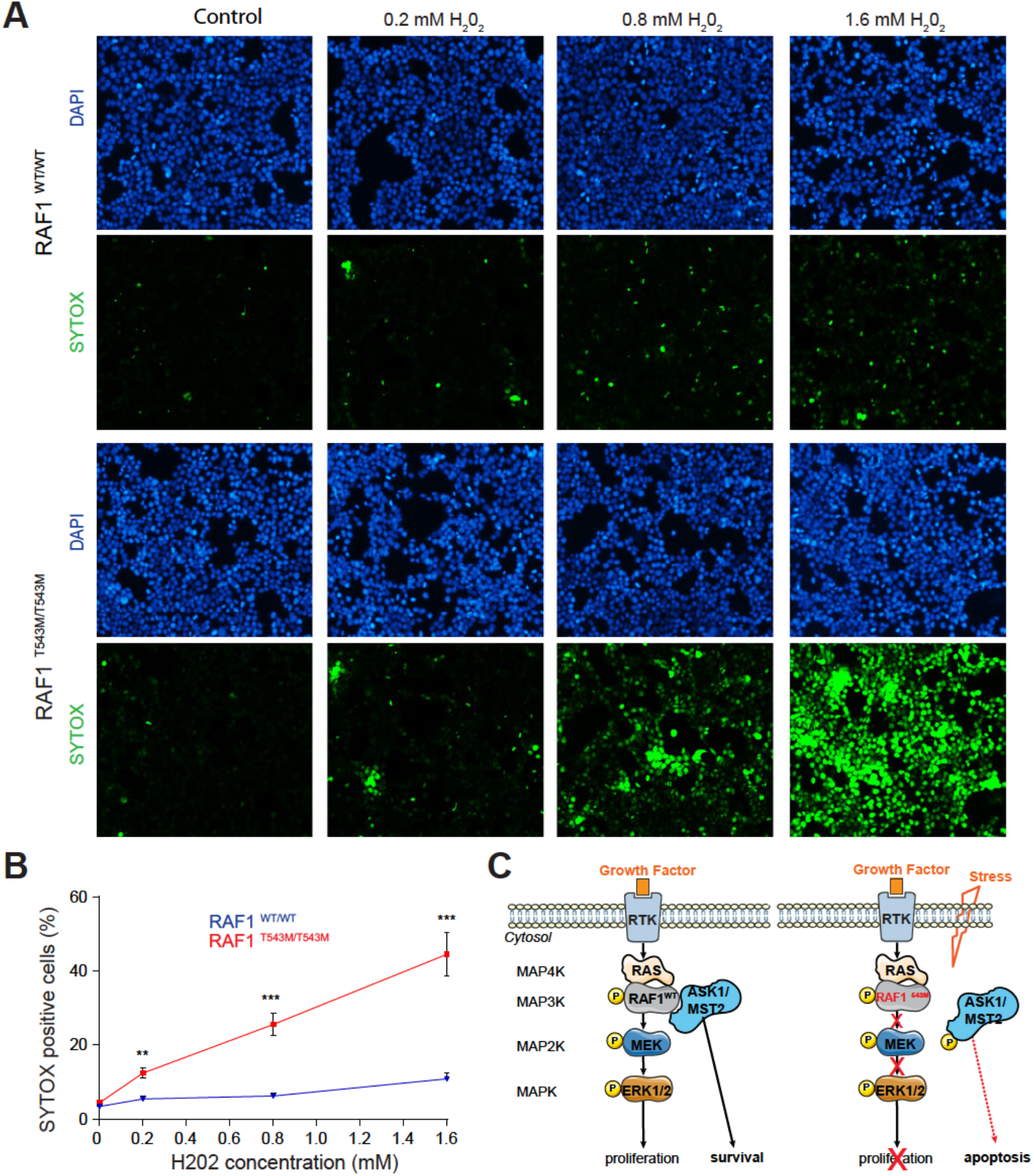
RAF1^T543M/T543M^ knock-in cells are more sensitive to stress-induced apoptosis. **(A)** Representative immunofluorescence images of 0.2, 0.8 and 1.6 mM H_2_O_2_ treated RAF1^T543M/T543M^ and RAF1^WT/WT^ HEK293T cells. Green: SYTOX dead cell stain. Blue: DAPI nuclei stain. Scale bar, 100 μm. **(A)** Dose-dependent SYTOX signal of RAF1^T543M/T543M^ and RAF1^WT/WT^ HEK293T cells incubated with increasing H_2_O_2_ concentrations for 2 hours. (Error bars indicate mean ± SEM. Two-tailed Student’s t test, **p<0.01, ***p<0.001). **(C)** Proposed model of RAF1^T543M/T543M^’s impact on proliferation via reduced MAPK/ERK transduction and increased apoptosis via derepression of ASK1/MST2 signaling.

## DISCUSSION

Collectively, our data unveil the genetic and molecular etiology of a novel segmental progeroid syndrome and reveal the role of wildtype RAF1 during human development and aging.

Our lines of investigations have highlighted several ways by which this germline RAF1^T543M^ mutation may behave as a strong hypomorphic allele which results in the neonatal lethal syndrome described herein. Our functional tests suggest that this RAF1^T543M^ variant: 1) is not actively phosphorylated at key activating residues including Ser259 and Ser338, 2) is unable *in vitro* and *in vivo*, to transduce RAS-mediated MAPK signaling towards MEK/ERK substrates, 3) is inherently unstable and prone to proteasome-mediated degradation and 4) is unable to block stress-induced apoptosis.

RAF1’s kinase activity is intrinsically linked to its own phosphorylation status. Tyrosine kinases such as SRC6, serine/threonine kinases such as CK2, and RAF1 itself, have been shown to impact RAF1’s phosphorylation.^28^ Here, we provide evidence that the residue Tyr543 which is mutated in this consanguineous family is unlikely to regulate RAF1 activity in a phosphorylation-dependent manner as substitutions with phosphorylation-inert residues have no effect on RAF1 activity. Incidentally, in zebrafish, the corresponding Tyr539 residue is a phenylalanine, while in mice, the corresponding Thr543 is replaced by Alanine. Taken together, our results suggest that the evolutionary changes at this residue in humans did not involve the creation of an additional regulatory phosphorylation site, rather we contend that the identified p.T543M missense mutation exerts its pathogenicity through an alternative mechanism, involving deficient kinase activity and reduced protein stability.

Apart from the established role of RAF1 in MAPK signaling, it has also been found to perform several kinase-independent functions. These include inhibition of ROCK2^29^ or promotion of cell survival by antagonizing proapoptotic kinases ASK1^10^ and MST2^27^. Here, we observed that the RAF1^T543M^ protein is markedly unstable compared to its wildtype counterpart. This instability significantly heightens cellular sensitivity to redox stress-induced apoptosis, which could be due to the loss of RAF1-mediated repression of proapoptotic kinases like ASK1 and MST2.

Given the central role played by RAF1 in transducing the cell growth promoting activities of numerous mitogens, it is fitting that a loss-of-function RAF1 mutant allele would have such consequences, especially during embryogenesis when the organism experiences exponential growth. The developmental syndrome described herein is reminiscent of Acro-cardio-facial syndrome (ACFS, MIM600460). ACFS is a very rare, genetically-orphan disorder characterized by split-hand/split-foot malformations (SHFM), congenital heart defects (CHD), facial anomalies, cleft lip/palate, and genital anomalies. First described by Richieri-Costa and Orquizas in 1987, a total of eight patients from six unrelated families have been reported.^30–35^ This syndrome is associated with very poor life expectancy, most patients surviving only a few hours or months. To date, no genetic cause has been established, although an autosomal-recessive inheritance pattern is supported by both consanguinity and recurrence in sibs born to unaffected parents.^34–36^

Notably, the cutaneous, cardiac and limb anomalies of the two affected siblings we described above are similar to those documented in *Raf1* mutant animal models. Wojnowski and colleagues noted thinner and less differentiated dermal and epidermal layers in *Raf1-/-* mice^9^, while Yamaguchi and colleagues found that heart-specific *Raf1-/-* mice exhibited cardiac dysfunction and apoptosis.^10^ More recently, the recessive-acting *wingless2* mutation found in *Gallus gallus* – which results in a phenotype strikingly similar to this syndrome with truncated limbs, craniofacial, cardiac and skin/feather defects – was linked to a homozygous premature stop codon in *Raf1*.^37^

These observations lead us to conjecture that the Mendelian disease and by inference ACFS may be caused by impaired RAF1 signaling. Hence, it will be worthwhile checking for mutations in RAF1, its upstream regulators and downstream effectors in previously reported ACFS cases. While we propose RAF1 insufficiency as a plausible genetic aetiology for ACFS, we contend that this does not imply that it should be regarded as a RASopathy. In contrast to the more common gain-of-function RAF1 mutations seen in Noonan syndrome and other RASopathies, ACFS reflects instead on the physiological requirement of wild type RAF1, the proto-oncogene rather than the oncogene. Like for other progeroid conditions such as *PYCR1* inactivation in De Barsy Syndrome, this RAF1 loss-of-function phenotype provides another illustration of how genes that are hijacked in the context of cancer may inversely cause premature aging when inactivated.^38,39^

## Data Availability

All data produced in the present study are available upon reasonable request to the authors.

## Non-standard abbreviations

CNS: Central nervous system
DCM: Dilated cardiomyopathy
ECHO: Echocardiogram
EGF: Epidermal growth factor
FGF(R): Fibroblast growth factor (receptor)
GoF: Gain-of-function
HCM: Hypertrophic cardiomyopathy
HDR: Homology-directed repair
LoF: Loss-of-function
OFC: Occipital frontal circumference
PDA: Patent ductus arteriosus
PFO: Patent foramen ovale
TRK: receptor tyrosine kinases
USG: Ultrasonography
VSD: Ventricular septal defect

## Author Contributions

NEB and BR directed the project and designed the experiments. AL and YXT performed genetic validation. SW performed *in vitro* assay, phosphorylation and *Xenopus* work. YXT generated and characterized RAF1 mutants and knock-in HEK293T cells. KYT and AL conducted viability assay. HK diagnosed and provided clinical information about the 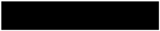 family. ZA and EÖ generated the predicted structural RAF1 mutant analyses. SW, YXT, NEB and BR wrote the manuscript with input from HK.

## Acknowledgements

We are grateful to all the individuals and the family for their participation in this research. We are grateful to all members of the Reversade laboratory for support and constructive feedback. BR was funded by a Strategic Positioning Fund for Genetic Orphan Diseases, an inaugural A*STAR Investigatorship from the Agency for Science, Technology and Research in Singapore, and is a fellow of the Branco Weiss Foundation and a National Research Foundation and EMBO Young Investigator.

## EXPERIMENTAL MODEL AND SUBJECT DETAILS

### Human Subjects

Peripheral blood samples were collected from 6 family members (II:1, II:3, III:2, III:4, III:5, III:6). Blood DNA was extracted using standard methods. All human studies were reviewed and approved by the Turkish and Singaporean institutional review board (A*STAR IRB #2019-087 and Koç University 2015.120.IRB2.047). Parents of the patient gave informed consent to publish their deceased child photographs.

## METHOD DETAILS

### Genetic analysis

SNP and linkage analysis were carried out on 5 family members (II:2, II:3, III:2, III:4 and III:5) using Affymetrix GeneChip Human Mapping 250K SNP microarray. Total of 5 family members were genotyped using Illumina HumanCore-12v1 BeadChips following manufacturer’s instructions. Call rates were above 99%, gender and relationship were 23 verified using Illumina GenomeStudio software. Linkage analysis was performed by searching for shared regions in the affected individual using custom programs written in Mathematica (Wolfram Research, Inc.). Whole-exome sequencing was performed on DNA from proband III:2. Briefly, the exome library was prepared on an Ion OneTouch System and sequenced on an Ion Proton instrument (Life Technologies) with one Ion PI chip. Sequence reads were aligned to the human GRCh37/hg19 assembly (UCSC Genome Browser). Variants were filtered for common SNPs against the NCBI’s ‘‘common and no known medical impacts’’ database (ClinVar), the Exome Aggregation Consortium (ExAC) Browser, the NHLBI Exome Sequencing Project, and an in-house database of 197 sequenced samples. Sanger sequencing was performed using primers flanking the mutations (Forward 5’-TAATGAAAGGGACAGCCTGG and Reverse 5’- CTCCCACCTTATATTGCCATC).

### Cell culture

HEK293T wildtype and mutant were grown in Dulbecco’s modified Eagle’s medium (DMEM) with 10% Fetal Bovine Serum (FBS, Hyclone), 1% L-glutamine (Invitrogen) at 37ºC in a humidified atmosphere of 5% CO2 plates. At various time points, cells were either fixed in 4% paraformaldehyde for further immunofluorescence analysis or collected for protein extraction.

### RAF1 overexpression assay *in vitro*

Full-length RAF1 cDNA (gift from Dhandapany’s group) was cloned into pCS2+ plasmid backbone and site-directed mutagenesis (QuikChange Site-Directed Mutagenesis kit, Stratagene) performed. Prior to transfection, HEK293T cells were plated onto poly-L-lysine (Sigma) pre-coated 6-well plate. At ∼90% confluence, cells were transfected with constructs using Lipofectamine 2000 (Invitrogen). Amounts of each construct transfected were adjusted to approximate equal RAF1 levels at collection. 48 hours after transfection, cells were serum-starved for 16 hrs by replacing with FBS-free media. Cells were then exposed for 15 min to FBS-containing media to initiate MAPK signalling, and harvested immediately on ice, for 30 min, using cold RIPA buffer supplemented with protease inhibitor cocktail (Roche) and phosphatase inhibitor (Sigma). Lysates were centrifuged at 4ºC, and supernatant collected for analysis.

### Antibodies

All specific antibodies were used between 1 and 10 µg/ml for western blotting and are commercially available: α-ACTIN (Chemicon, Mab1501R), c-RAF (BD Biosciences, 610152), phospho-c-RAF(Ser259) (Cell Signaling, 9421), phospho-c-RAF (Ser338) (Cell Signaling, 9427), pMEK1/2 (Cell Signaling, 9154), total MEK1/2 (Cell Signaling, 4694), pERK1/2 (Cell Signaling, 9101), total ERK1/2 (Cell Signaling, 9102).

### RAF1 mutant cells generation

An sgRNA against exon 15 of RAF1 (20 bp sgRNA sequence: 5’-ATTGTATGAACTGATGACGG-3’) was cloned into a plasmid for coexpression with SpCas9-T2A-Puro (pSpCas9(BB)-2A-Puro (PX459) V2.0, Addgene plasmid # 62988) according to Ran et al. 2013^40^. This sgRNA was chosen as it contains the target c.1628C nucleotide 1 bp from the Cas9 cut site, for maximal substitution efficiency. ^41^ For HDR, a single-stranded oligodeoxynucleotide (ssODN) with 40 bp homology arms flanking the desired c.1628C>T (p.T543M) and c.1632G>C (PAM disruption, synonymous) mutations was designed. Successful c.1628C>T HDR would create a BccI restriction site. ssODN sequence:

5’- GATGTCTACTCCTATGGCATCGTATTGTATGAACTGATGATGGGCGAGCTTCCTTATTCTCACATCAACAACCGAGATCAG-3’

A 70% confluent 12-well plate of HEK293T cells was transfected with 625ng RAF1 sgRNA+PX459 and 125 µl of 10 µM ssODN using Lipofectamine 2000 (Life technologies), following manufacturer’s protocol. 24 hours post-transfection, media was replaced with puromycin to a final concentration of 2 µg/ml for 4 days selection. Surviving cells were then serially diluted and plated into 96-well plate at 0.5 cells/well for clonal expansion. Wells with a single colony were Sanger-sequenced during splitting and successful HDR confirmed by PCR and BccI restriction digest. Due to the low frequency of HDR, the majority of clones contained indels (39/43 clones analysed), with a single compound heterozygous 2 bp indel/c.1628C>T and a single homozygous c.1628C>T clone obtained. A clone with homozygous 17 bp indels was retained as RAF1^KO/KO^ cells. The compound heterozygote was verified by TA-cloning RAF1 exons 15-16, and subsequently Sanger-sequencing individual transformed colonies, yielding a ∼1:1 mix of two alleles (data not shown). pSpCas9(BB)-2A-Puro (PX459) V2.0 was a gift from Feng Zhang (Addgene plasmid #62988; http://n2t.net/addgene:62988 ; RRID:Addgene_62988).

### Cell viability assay

Wild-type or knock-in HEK293T cells were plated in 12-well plates pre-coated with poly-L-lysine (Sigma). Upon confluence, cells were stimulated with a range of H2O2 (0.2, 0.8 or 1.6 mM) and further incubated at 37°C for 2 hours. H2O2 concentrations were carefully chosen to induce minimum apoptosis in HEK293T wildtype cells. Cell death was assessed by adding SYTOX® Green Nucleic Acid Stain (1/1000, Invitrogen, S7020) in the culture for 15 min. Subsequently, cells were incubated with DAPI. Images were taken with a Thermo Fisher Scientific EVOS® FL Cell Imaging System. SYTOX green and DAPI blue positive nuclei were counted automatically using ImageJ and data were expressed in percentage per number total of nuclei. Two independent assays were performed and 4 representative images were analyzed for each condition.

### Biochemistry

For western blotting, cells or *Xenopus* embryos were lysed in RIPA buffer supplemented with protease inhibitor cocktails (Roche) and phosphatase inhibitors (Sigma). Extracted proteins were separated by electrophoresis on SDS polyacrylamide gels with DTT (Bio-Rad), followed by a TransBlot Turbo transfer (Bio-Rad) onto polyvinylidene difluoride (PVDF) membranes. Membranes were incubated with primary antibodies in 5% BSA or 5% milk in Tris-Buffered Saline-Tween (TBST) according to the manufacturer’s instructions for 2 hours at room temperature or 4°C overnight on an orbital shaker, followed by three washes of 15 min in TBST. Following a 2-hour incubation with the respective secondary antibodies, membranes were washed in TBST and developed using West Pico, Dura or Femto chemiluminescence reagents (Thermo Fisher). Global phosphorylation was measured using a 7.5% SuperSep™ Phos-tag™ gel. Protein samples were run in Tris-Glycine SDS buffer according to the manufacturer’s instructions followed by a wet transfer method to a PVDF membrane in 20% methanol Tris-Glycine SDS buffer. The membrane was immunoblotted in the same manner as described for Western blots above. To block protein ubiquitin-dependent degradation 100 nM epoxomicin (Sigma) was added to cultured cells for 8 or 24 hours. Quantification of western blot was performed by ImageJ.

### *Xenopus* methods

Protocols for *Xenopus* fertilization, micro-injections, mRNA synthesis, and whole-mount in situ hybridization (WISH) are on our protocol’s website page (see http://www.reversade.com-a.googlepages.com/protocols/). 200 pg of either RAF1^WT^, RAF1^T543M^, RAF1^L603P^ or RAF1^S257L^ mRNA were injected into 4-cell stage *Xenopus laevis* embryos, and harvested at various developmental stages for subsequent protein extraction or WISH analyses. Images of embryos were captured with a stereomicroscope equipped with an integrated digital camera (Leica; M205 FA).

### Quantification and statistical analysis

All statistical tests were carried out using Prism 7 or Excel unless otherwise stated. Information on statistical tests used for each assay and number of samples are detailed in the Figure legends and in Method sections.

## Supplemental Data

**Figure S1.**
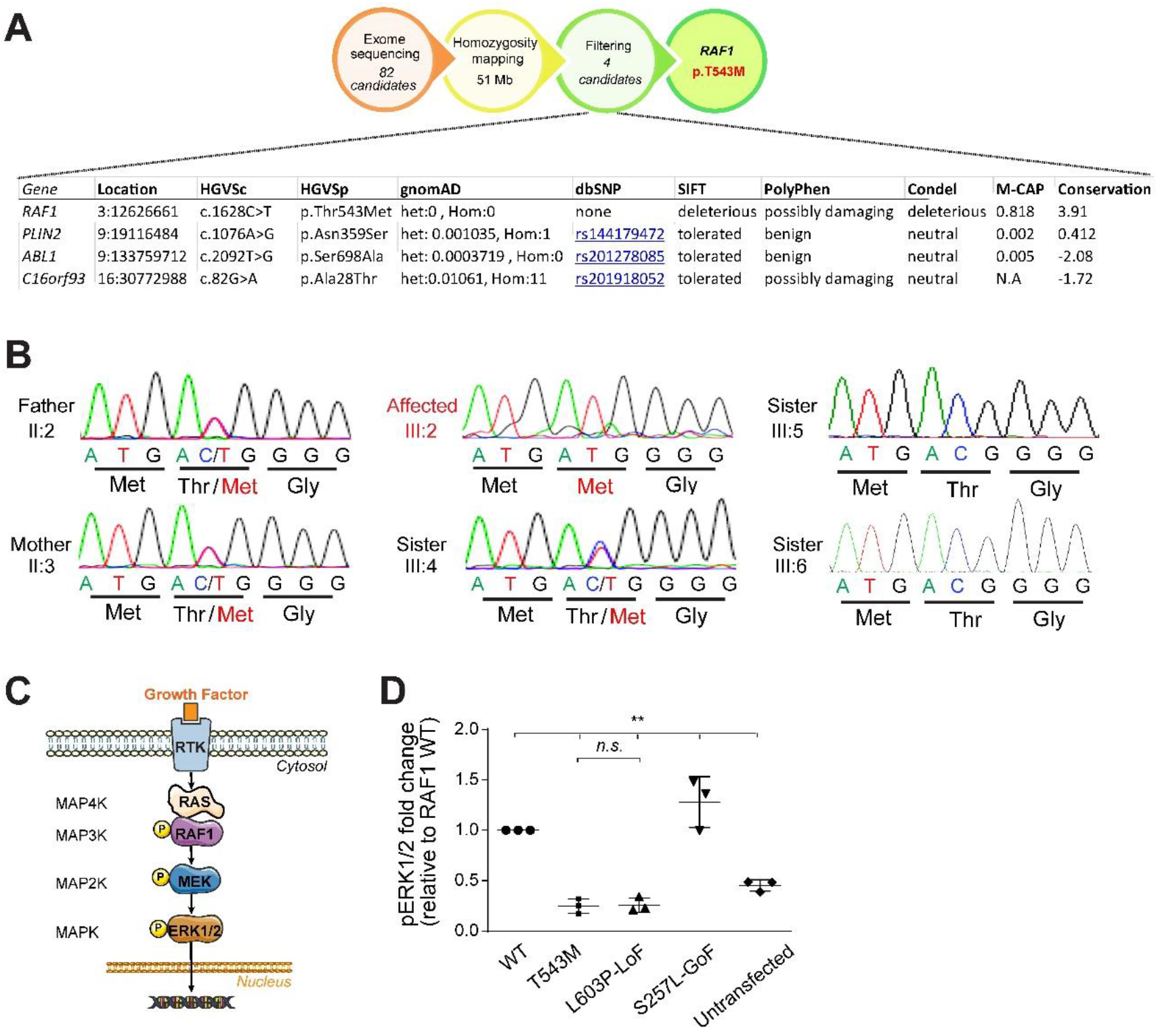
Related to Figure 1. **(A)** Genetic strategy. Combination of exome sequencing and homozygosity mapping. resulted in 4 candidate genes. Germline homozygous variants in *PLIN2, ABL1* and *C16orf93* were filtered out due to their low phylogenetic conservation and low predicted deleterious potential. **(B)** Sanger sequencing chromatogram showing variant c.1628C>T (p.T543M) segregating with disease: homozygous in the proband (III:2), heterozygous in the parents (II:2 and II:3) and one sister (III:4) and absent in two other siblings (III:3 and III:6). **(C)** Schematic representation of ERK1/2 MAP Kinase pathway. **(D)** Quantification of phosphorylated ERK1/2, normalized to WT RAF1 in 3 independent western blots (Error bars indicate mean ± SEM. Two-tailed Student’s t test. *n*.*s*. not significant, **p<0.01).

**Figure S2.**
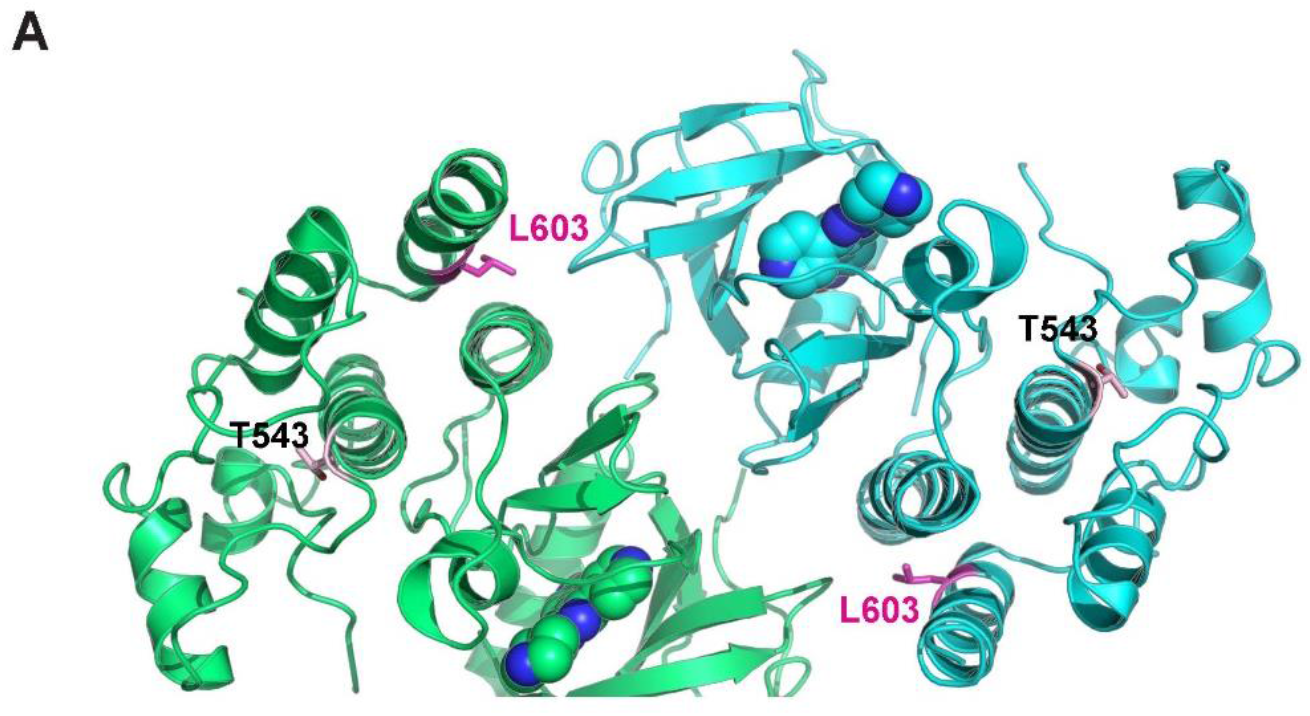
Related to Figure 3. Position of Leu603 and Thr543 in the RAF1 homodimer (PDB:3OMV). The p.L603P mutation causes an impairment of kinase activity, likely through destabilizing the αH helix and the helical bundle made by the αC and αH helices, and may also affect the dimer stability. Thr543 is located in the αE helix.

## References

1. Leicht, D.T., Balan, V., Kaplun, A., Singh-Gupta, V., Kaplun, L., Dobson, M., and Tzivion, G. (2007). Raf kinases: Function, regulation and role in human cancer. Biochimica et Biophysica Acta - Molecular Cell Research 1773, 1196–1212.

2. Rauen, K.A. (2013). The RASopathies. Annu. Rev. Genomics Hum. Genet. 14, 355–369.

3. Baccarini, M. (2005). Second nature: Biological functions of the Raf-1 “kinase.” FEBS Lett. 579, 3271–3277.

4. Motta, M., Fasano, G., Gredy, S., Brinkmann, J., Bonnard, A.A., Simsek-Kiper, P.O., Gulec, E.Y., Essaddam, L., Utine, G.E., Guarnetti Prandi, I., et al. (2021). SPRED2 loss-of-function causes a recessive Noonan syndrome-like phenotype. Am. J. Hum. Genet. 108, 2112–2129.

5. Aoki, Y., Niihori, T., Inoue, S.-I., and Matsubara, Y. (2016). Recent advances in RASopathies. J. Hum. Genet. 61, 33–39.

6. Tajan, M., Paccoud, R., Branka, S., Edouard, T., and Yart, A. (2018). The RASopathy family: Consequences of germline activation of the RAS/MAPK pathway. Endocr. Rev. 39, 676–700.

7. Pritchard, C.A., Bolin, L., Slattery, R., Murray, R., and McMahon, M. (1996). Post-natal lethality and neurological and gastrointestinal defects in mice with targeted disruption of the A-Raf protein kinase gene. Curr. Biol. 6, 614–617.

8. Wojnowski, L., Zimmer, A.M., Beck, T.W., Hahn, H., Bernal, R., Rapp, U.R., and Zimmer, (1997). Endothelial apoptosis in braf-deficient mice. Nat. Genet. 16, 293–297.

9. Wojnowski, L., Stancato, L.F., Zimmer, A.M., Hahn, H., Beck, T.W., Larner, A.C., R. Rapp U., and Zimmer, A. (1998). Craf-1 protein kinase is essential for mouse development. Mech. Dev. 76, 141–149.

10. Yamaguchi, O., Watanabe, T., Nishida, K., Kashiwase, K., Higuchi, Y., Takeda, T., Hikoso, S., Hirotani, S., Asahi, M., Taniike, M., et al. (2004). Cardiac-specific disruption of the c-raf-1 gene induces cardiac dysfunction and apoptosis. J. Clin. Invest. 114, 937–943.

11. Dumaz, N., and Marais, R. (2005). Raf phosphorylation: one step forward and two steps back. Mol. Cell 17, 164–166.

12. Dhillon, A.S., Meikle, S., Yazici, Z., Eulitz, M., and Kolch, W. (2002). Regulation of Raf-1 activation and signalling by dephosphorylation. EMBO J. 21, 64–71.

13. Chen, J., Fujii, K., Zhang, L., Roberts, T., and Fu, H. (2001). Raf-1 promotes cell survival by antagonizing apoptosis signal-regulating kinase 1 through a MEK-ERK independent mechanism. Proceedings of the National Academy of Sciences 98, 7783–7788.

14. Dhandapany, P.S., Razzaque, M.A., Muthusami, U., Kunnoth, S., Edwards, J.J., Mulero-Navarro, S., Riess, I., Pardo, S., Sheng, J., Rani, D.S., et al. (2014). RAF1 mutations in childhood-onset dilated cardiomyopathy. Nat. Genet. 46, 635–639.

15. Jaffré, F., Miller, C.L., Schänzer, A., Evans, T., Roberts, A.E., Hahn, A., and Kontaridis, M.I. (2019). Inducible Pluripotent Stem Cell-Derived Cardiomyocytes Reveal Aberrant Extracellular Regulated Kinase 5 and Mitogen-Activated Protein Kinase Kinase 1/2 Signaling Concomitantly Promote Hypertrophic Cardiomyopathy in RAF1-Associated Noonan Syndrome. Circulation 140, 207–224.

16. Umbhauer, M., Marshall, C.J., Mason, C.S., Old, R.W., and Smith, J.C. (1995). Mesoderm induction in Xenopus caused by activation of MAP kinase. Nature 376, 58–62.

17. MacNicol, A.M., Muslin, A.J., and Williams, L.T. (1993). Raf-1 kinase is essential for early Xenopus development and mediates the induction of mesoderm by FGF. Cell 73, 571–583.

18. Delaune, E., Lemaire, P., and Kodjabachian, L. (2005). Neural induction in Xenopus requires early FGF signalling in addition to BMP inhibition. Development 132, 299–310.

19. Pera, E.M., Wessely, O., Li, S.Y., and De Robertis, E.M. (2001). Neural and head induction by insulin-like growth factor signals. Dev. Cell 1, 655–665.

20. Hatzivassiliou, G., Song, K., Yen, I., Brandhuber, B.J., Anderson, D.J., Alvarado, R., Ludlam, M.J.C., Stokoe, D., Gloor, S.L., Vigers, G., et al. (2010). RAF inhibitors prime wild-type RAF to activate the MAPK pathway and enhance growth. Nature 464, 431–435.

21. Noble, C., Mercer, K., Hussain, J., Carragher, L., Giblett, S., Hayward, R., Patterson, C., Marais, R., and Pritchard, C.A. (2008). CRAF autophosphorylation of serine 621 is required to prevent its proteasome-mediated degradation. Mol. Cell 31, 862–872.

22. Wu, X., Yin, J., Simpson, J., Kim, K.-H., Gu, S., Hong, J.H., Bayliss, P., Backx, P.H., Neel, B.G., and Araki, T. (2012). Increased BRAF Heterodimerization Is the Common Pathogenic Mechanism for Noonan Syndrome-Associated RAF1 Mutants. Mol. Cell. Biol. 32, 3872–3890.

23. Hindley, A., and Kolch, W. (2002). Extracellular signal regulated kinase (ERK)/mitogen activated protein kinase (MAPK)-independent functions of Raf kinases. J. Cell Sci. 115, 1575–1581.

24. Desideri, E., Cavallo, A.L., and Baccarini, M. (2015). Alike but Different: RAF Paralogs and Their Signaling Outputs. Cell 161, 967–970.

25. Xiang, J., Wan, C., Guo, R., and Guo, D. (2016). Is Hydrogen Peroxide a Suitable Apoptosis Inducer for All Cell Types? BioMed Research International 2016, 1–6.

26. Romano, D., Matallanas, D., Weitsman, G., Preisinger, C., Ng, T., and Kolch, W. (2010). Proapoptotic kinase MST2 coordinates signaling crosstalk between RASSF1A, Raf-1, and Akt. Cancer Res. 70, 1195–1203.

27. O’Neill, E., Rushworth, L., Baccarini, M., and Kolch, W. (2004). Role of the kinase MST2 in suppression of apoptosis by the proto-oncogene product Raf-1. Science 306, 2267–2270.

28. Matallanas, D., Birtwistle, M., Romano, D., Zebisch, A., Rauch, J., von Kriegsheim, A., and Kolch, W. (2011). Raf family kinases: old dogs have learned new tricks. Genes Cancer 2, 232–260.

29. Ehrenreiter, K., Piazzolla, D., Velamoor, V., Sobczak, I., Small, J.V., Takeda, J., Leung, T., and Baccarini, M. (2005). Raf-1 regulates Rho signaling and cell migration. J. Cell Biol. 168, 955–964.

30. Guion-Almeida, M.L., Zechi-Ceide, R.M., and Richieri-Costa, A. (2000). Cleft lip/palate, abnormal ears, ectrodactyly, congenital heart defect, and growth retardation: definition of the acro-cardio-facial syndrome. Clin. Dysmorphol. 9, 269–272.

31. Mingarelli, R., Zuccarello, D., Digilio, M.C., and Dallapiccola, B. (2005). A new observation of acro-cardio-facial syndrome substantiates interindividual clinical variability. Am. J. Med. Genet. A 136, 84–86.

32. Sivasli, O., Ozer, E.A., Ozer, A., Aydinlioglu, H., and Helvaci, M. (2007). Acro-cardio-facial syndrome associated with neuroepithelial cyst: a case report. Genet. Couns. 18, 247–250.

33. Tanpaiboon, P., Sittiwangkul, R., Dejkhamron, P., Srikummool, M., Sripathomsawat, W., and Kantaputra, P. (2009). Expanding the phenotypic spectrum of acro-cardio-facial syndrome (ACFS): Exclusion of P63 mutation. Am. J. Med. Genet. A 149, 1749–1753.

34. Giannotti, A., Digilio, M.C., Mingarelli, R., and Dallapiccola, B. (1995). An autosomal recessive syndrome of cleft palate, cardiac defect, genital anomalies, and ectrodactyly (CCGE). J. Med. Genet. 32, 72–74.

35. Kariminejad, A., Bozorgmehr, B., Sedighi Gilani, M.A., Almadani, N., and Kariminejad, M.H. (2008). Clinical variability in acro-cardio-facial-syndrome. Am. J. Med. Genet. A 146A, 1977–1979.

36. Richieri Costa, A., and Orquizas, L.C. (1987). Ectrodactyly, cleft lip/palate, ventricular septal defect, micropenis and mental retardation in a brazilian child born to consaguineous parents. Rev. bras. genét, 787–792.

37. Youngworth, I., and Delany, M.E. (2019). A Premature Stop Codon in RAF1 Is the Priority Candidate Causative Mutation of the Inherited Chicken Wingless-2 Developmental Syndrome. Genes 10.

38. Ding, Z., Ericksen, R.E., Escande-Beillard, N., Lee, Q.Y., Loh, A., Denil, S., Steckel, M., Haegebarth, A., Wai Ho, T.S., Chow, P., et al. (2020). Metabolic pathway analyses identify proline biosynthesis pathway as a promoter of liver tumorigenesis. J. Hepatol. 72, 725–735.

39. Reversade, B., Escande-Beillard, N., Dimopoulou, A., Fischer, B., Chng, S.C., Li, Y., Shboul, M., Tham, P.-Y., Kayserili, H., Al-Gazali, L., et al. (2009). Mutations in PYCR1 cause cutis laxa with progeroid features. Nat. Genet. 41, 1016–1021.

40. Ran, F.A., Hsu, P.D., Lin, C.-Y., Gootenberg, J.S., Konermann, S., Trevino, A.E., Scott, D.A., Inoue, A., Matoba, S., Zhang, Y., et al. (2013). Double nicking by RNA-guided CRISPR Cas9 for enhanced genome editing specificity. Cell 154, 1380–1389.

41. Paquet, D., Kwart, D., Chen, A., Sproul, A., Jacob, S., Teo, S., Olsen, K.M., Gregg, A., Noggle, S., and Tessier-Lavigne, M. (2016). Efficient introduction of specific homozygous and heterozygous mutations using CRISPR/Cas9. Nature 533, 125–129.

